# Multi-Omic Profiling of Plasma Identify Biomarkers and Pathogenesis of COVID-19 in Children

**DOI:** 10.1101/2021.03.04.21252876

**Authors:** Chong Wang, Xufang Li, Wanshan Ning, Sitang Gong, Fengxia Yang, Chunxiao Fang, Yu Gong, Di Wu, Muhan Huang, Yujie Gou, Shanshan Fu, Yujie Ren, Ruyi Yang, Yang Qiu, Yu Xue, Yi Xu, Xi Zhou

**Affiliations:** Guangzhou Institute of Pediatrics, Guangzhou Women and Children’s Medical Center, Guangzhou, Guangdong, 510120, China; State Key Laboratory of Virology, Wuhan Institute of Virology, Center for Biosafety Mega-Science, Chinese Academy Sciences, Wuhan, Hubei 430071, China; MOE Key Laboratory of Molecular Biophysics, Hubei Bioinformatics and Molecular Imaging Key Laboratory, Center for Artificial Intelligence Biology, College of Life Science and Technology, Huazhong University of Science and Technology, Wuhan, Hubei 430074, China

## Abstract

Although children usually develop less severe disease responding to COVID-19 than adults, little is known about the pathogenesis of COVID-19 in children. Herein, we conducted the plasma proteomic and metabolomic profiling of a cohort of COVID-19 pediatric patients with mild symptoms. Our data show that numerous proteins and metabolites involved in immune as well as anti-inflammatory processes were up-regulated on a larger scale in children than in adults. By developing a machine learning-based pipeline, we prioritized two sets of biomarker combinations, and identified 5 proteins and 5 metabolites as potential children-specific COVID-19 biomarkers. Further study showed that these identified metabolites not only inhibited the expression of pro-inflammatory factors, but also suppressed coronaviral replication, implying that these factors played key roles in protecting pediatric patients from both viral infection and infection-induced inflammation. Together, our study uncovered a protective mechanism responding to COVID-19 in children, and sheds light on potential therapies.

**Teaser:** Anti-inflammatory metabolites were highly elevated in the plasma of COVID-19 pediatric patients with mild symptoms.

## MAIN TEXT

### Introduction

The pandemic of Coronavirus Disease 2019 (COVID-19) caused by severe acute respiratory syndrome coronavirus 2 (SARS-CoV-2) has become the worst public health crisis once a century, which has caused over 97 million human infections and 2 million deaths all over the world. It had been found that all people are susceptible to SARS-CoV-2 without significant differences in sex or age ^1-3^, and SARS-CoV-2 infects children under 18-year-old at a similar rate as adults^4^. Reports from different countries showed that the symptoms are milder in the overwhelming majority of children with COVID-19 (CC) compared to that of adults with COVID-19 (AC)^1-3, 5-11^. Most children have minor symptoms or an asymptomatic SARSCoV2 infection and severe conditions such as acute respiratory distress syndrome and multisystem inflammatory syndrome are rare in CC^12-14^.

Several theories have been discussed to explain the differences in clinical symptoms between CC and AC^15^. One possible theory is the differences in the composition and functional responsiveness of immune systems between children and adults^16^. Children have a qualitatively different response to SARS-CoV-2 compared to adults^17^. Besides, young children were usually infected with other simultaneous viruses in the mucosa of lungs and airways^18^, which would restrict the infection of SARS-CoV-2 via virus to virus interactions and competition. Another possible explanation is the differences in the maturity and function of the viral entry receptor angiotensin-converting enzyme (ACE2) between children and adults. In addition, children have the better overall physical conditions compared to adults, which probably make children more resistant for SARS-CoV-2^19^. However, the detailed mechanisms for the differences in clinical symptoms between children and adults remain to be determined.

Multiple reports showed that immune system of CC is less likely to elicit an excessive inflammatory response and cytokine storm, as observed in AC, suggesting that global molecular alterations in CC might be much milder and the deterioration process of COVID-19 was not strongly induced in CC. Considering that the intrinsic differences between children and adults, we proposed that molecules associated with COVID-19 in CC were also induced, whereas some protective mechanisms were elicited to antagonize the deterioration of the disease. To test this hypothesis, we collected plasma samples from a cohort including 18 CC cases and 12 healthy children (HC), and conducted the proteomic and metabolomic profiling. By comparing the omics data of CC with those of adult with COVID-19 (AC) that identified previously by us^20, 21^, we uncovered numerous molecular alterations in CC against AC cases, which may contribute to the pathogenesis of COVID-19 in children. Moreover, we developed a new pipeline named inference of biomarker combinations with minimal bias (iBM), and predicted 5 proteins and 5 metabolites as potential CC-specific biomarkers. In addition, some of metabolite biomarkers were experimentally examined their roles in suppressing viral replication and/or modulating inflammation *in vitro*. Our findings provided valuable knowledge about plasma biomarkers associated with CC, shed lights on the better understanding of mild COVID-19 symptom in children, and may reveal potential therapeutic targets.

## Results

### Study design and blood samples

We collected the blood samples of 30 children including 18 CC and 12 HC cases from Guangzhou Women and Children’s Medical Center (Fig. 1A). All CC cases were diagnosed as mild symptoms based on the Diagnosis and Treatment Protocol for Novel Coronavirus Pneumonia (6^th^ edition) of the National Health Commission of China^22^, and discharged from the hospital after recovery. No severe or critically ill CC cases were charged in our hospital (Table 1). The 12 HC cases, whose throat swab and serological tests were negative for SARS-CoV-2, were enrolled for comparison (Table S1). The clinical data of CC is shown in Table 1.

**Table 1.** Characteristics of COVID-19 pediatric patients.

| Children with COVID-19 | n=18 |
| --- | --- |
| Age-year |  |
| Median (IQR) | 7 (5, 12) |
| Sex-no.(%) |  |
| Female | 5 (27.78%) |
| Male | 13 (72.22%) |
| Throat swab for SARS-CoV-2 (days) |  |
| Median (IQR) | 6 (4, 14) |
| Sampling time from the disease onset (days) |  |
| Median (IQR) | 7 (10, 13) |
| Co-infection-no.(%) |  |
| Other viruses | ND |
| Bacteria | ND |
| Fungus | ND |
| Clinical outcome-no.(%) |  |
| Discharged | 18 (100%) |
| Blood routine- Median (IQR) |  |
| WBC ($\times 10^9/L$, normal range 5-12) | 6.90 (5.50, 8.55) |
| Lymphocyte ($\times 10^9/L$, normal range 1.5-4.8) | 2.60 (2.07, 4.19) |
| Neutrophil ($\times 10^9/L$, normal range 2-7.2) | 2.58 (2.15, 3.24) |
| CD19+ (cells/$\mu l$, normal range 90-660) | 396.35 (236.82, 800.53) |
| CD3+ (cells/$\mu l$, normal range 690-2540) | 1232.74 (967.60, 2305.85) |
| CD3+CD4+ (cells/$\mu l$, normal range 410-1590) | 655.44 (459.88, 1150.56) |
| CD3+CD8+ (cells/$\mu l$, normal range 190-1140) | 482.67 (386.66, 859.49) |
| NK (cells/$\mu l$, normal range 90-590) | 334.34 (194.04, 438.04) |
| Platelet ($10^9/L$, normal range 140-440) | 299.50 (263.00, 333.75) |
| RBC ($\times 10^{12}/L$, normal range 4-4.5) | 4.66 (4.37, 5.36) |
| Haemoglobin (g/L, normal range 105-145) | 124.50 (119.00, 141.75) |
| APTT (s, normal range 28-45) | 40.50 (37.40, 42.60) |
| PT (s, normal range 11-15) | 13.40 (13.15, 13.90) |
| D-dimer ($\mu g/mL$, normal range, 0-1.5 ) | 0.31 (0.26, 0.40) |
| Cytokines- Median (IQR) |  |
| IFN-γ (pg/mL, normal range 0-6.56) | 4.62 (3.97, 6.07) |
| IL-10 (pg/mL, normal range 0-8.14) | 1.98 (1.74, 2.91) |
| IL-12p70 (pg/mL, normal range 0-6.9) | 2.89 (1.77, 4.38) |
| IL-17A (pg/mL, normal range 0-3.71) | 5.98 (3.67, 8.76) |
| IL-1β (pg/mL, normal range 0-3.12) | 0.94 (0.59, 1.70) |
| IL-2 (pg/mL, normal range 0-5.03) | 4.33 (2.71, 5.01) |
| IL-22 (pg/mL, normal range 0-2.61) | 2.53 (1.66, 4.40) |
| IL-4 (pg/mL, normal range 0-4.62) | 3.37 (2.89, 5.42) |
| IL-5 (pg/mL, normal range 0-3.73) | 2.45 (1.94, 3.47) |
| IL-6 (pg/mL, normal range 0-8.88) | 3.84 (2.33, 6.58) |
| IL-8 (pg/mL, normal range 0-15.71) | 6.55 (4.37, 9.34) |
| TNF-α (pg/mL, normal range 0-5.35) | 5.35 (4.12, 6.95) |

**Figure 1.**
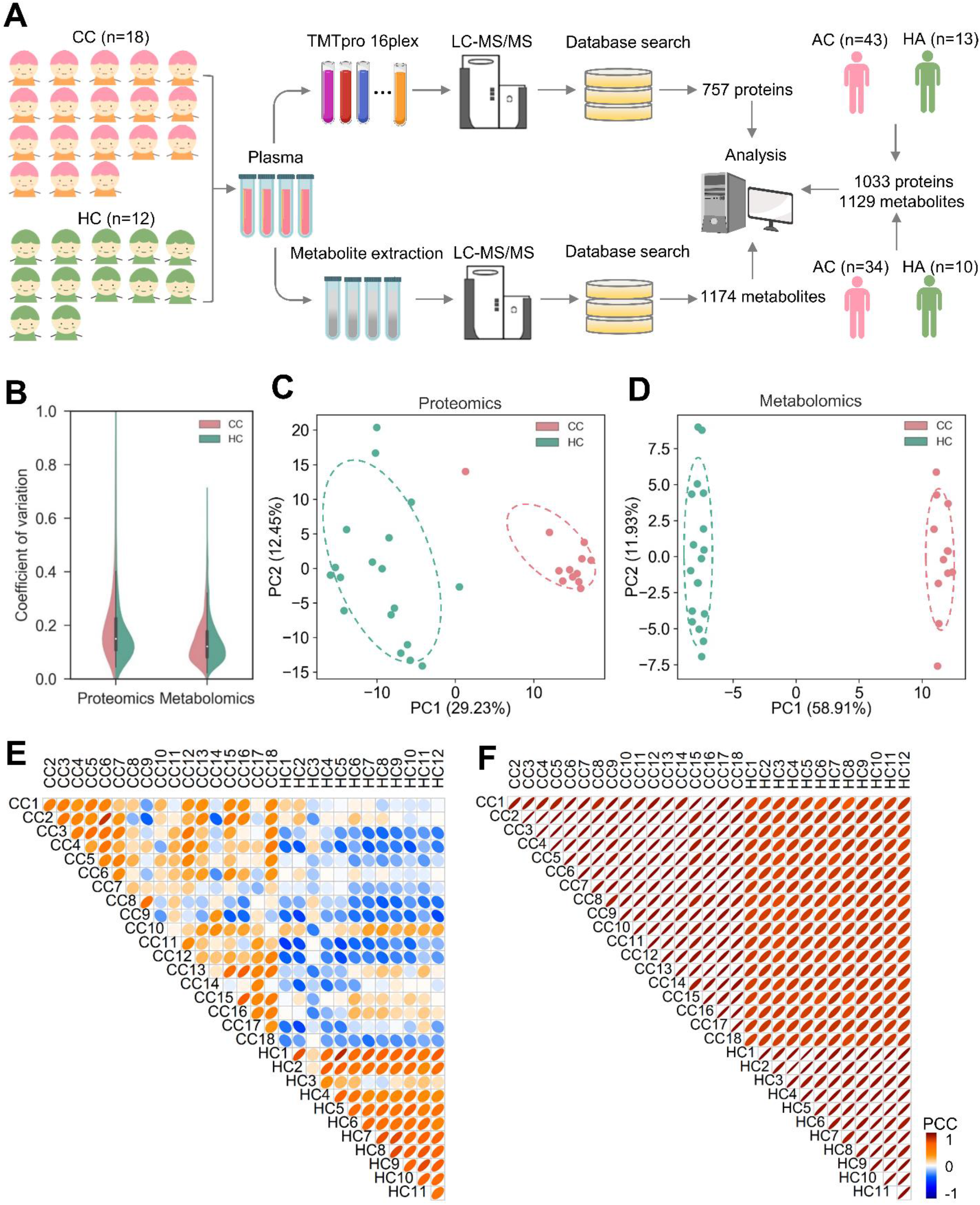
Study design and quality control of proteomic and metabolomic data. **A** Overview of plasma samples collection from COVID-19 children, including CC (*n* = 18) and HC (*n* = 12) cases. The workflow for processing the proteomic and metabolomic data was shown, including the plasma separation, TMTpro 16-plex labeling, metabolite extraction, LC-MS/MS analysis, database search and further computational analyses. The proteomic data of 43 AC and 13 HA cases, and metabolomic data of 34 AC and 10 HA cases from our two previous studies were also used for further computational analyses. **B** CV of the proteomic and metabolomic data. **C**,**D** The PCA analysis of the proteomic (C) and metabolomic (D) data. **E**,**F** PCC for each pair of samples using the proteomic (E) and metabolomic (F) data.

For each blood sample, plasma was separated, and total proteins were extracted, denatured and digested into peptides by trypsin (Fig. 1A). Then, the 30 plasma samples were separated into 2 batches and individually subjected to tandem mass tag (TMT) labeling (Table S2). For each batch, individual samples were labeled with TMT 16-plex reagents, and a pooling mixture of all the 30 samples was included and labeled as a standard control to eliminate the batch effect. After fractionation, each batch of peptide mixtures was analyzed by liquid chromatography with tandem mass spectrometry (LC-MS/MS). The hydrophilic and hydrophobic metabolites were extracted from each plasma sample, respectively and then determined via using liquid chromatography electrospray ionization tandem mass spectrometry (LC-ESI-MS/MS) system. The identification of metabolites was conducted by using home-made database with retention time and ion pairs or searching against the public databases based on MS/MS spectra for those metabolites without authentic standards in our database.

Recently, we conducted metabolomic and proteomic profilings of plasma samples from adults with or without SARS-CoV-2 infection, and identified numerous molecular alterations associated with COVID-19 in adults^20, 21^. From our two previous studies, we obtained the TMT-based quantitative proteomic data of 43 AC cases and 13 healthy adults (HA), and metabolomic data of 34 AC and 10 HA cases. In total, there were 1033 proteins and 1129 metabolites quantified from AC and HA cases.

### A multi-omic profiling of plasma samples

The proteomic profiling detected 9445 peptides from the 30 plasma samples, with an average number of 4716.1 peptides per sample (Fig. S1A). We mapped the peptides to corresponding protein sequences, and quantified proteins using the reporter ion MS2 module of the MaxQuant software package^23^. We found that 757 proteins were quantified in at least one sample (Table S3), with average numbers of 666 and 666 proteins in the 18 CC and 12 HC samples, respectively (Fig. S1B). We checked the raw MS/MS data and found that 4877 peptides (48.4%) could be matched by ≥ 2 spectral counts (Fig. S1C). The average spectral counts were calculated as 2.6 for all peptides, indicating a high quality of peptide identification. Also, we found that 626 proteins (79.2%) could be traced and supported by ≥ 2 peptides, with an average number of 11.1 peptides (Fig. S1D). Thus, our proteomic data was also highly reliable at the protein level. After the metabolomic profiling, from the principal component analysis (PCA) results, it was found that the 4 QC samples can be clustered in the center, indicating a high quality of metabolomic identification (Fig. S1E). We in total obtained 1174 metabolites from the 30 samples (Table S4), with an average number of 1140.7 metabolites per sample (Fig. S1F). The multiquant software package was used for quantification.

From the multi-omic profiling, the intensity-based abundances (IBAs) of proteins and metabolites were obtained. For each batch of the proteomic data, the relative protein abundances (RPA) were obtained by normalizing the control sample, and proteins not quantified in control were discarded. To ensure the data quality, only 575 proteins and 1158 metabolites mutually quantified in > 80% samples for further analysis, and a heatmap was illustrated after a hierarchical clustering (Fig. S2). It could be found that CC and HC cases have distinct molecular signatures. For each protein or metabolites, normal distribution imputation (NI) was applied to impute the missing values^24^. To measure the variability of proteomic and metabolomic quantification, the coefficient of variation (CV) was calculated for each molecule, with median values of 0.172 and 0.135 for the proteomic data, and 0.122 and 0.122 for the metabolomic data in CC and HC samples, respectively (Fig. 1B). The low CV values supported a high reproducibility of the multi-omic quantification. Then, the principal component analysis (PCA) was conducted for the proteomic (Fig. 1C) and metabolomic data (Fig. 1D). From the results, it was found that CC and HC cases could be clearly distinguished using either the data type, indicating molecular alterations were quite significant in both omic levels. In addition, we calculated the average Pearson correlation coefficient (PCC) for each pair of samples using the proteomic (Fig. 1E) or metabolomic data (Fig. 1F), and the results supported that similar molecular alterations triggered in CC cases.

### Characterization of CC-specific molecular alterations

To identify molecular alterations in CC against HC cases, we directly used RPAs of the proteomic data and IBAs of the metabolomic data, and in total detected 121 and 418 potential DEPs and DEMs, respectively (Fig. 2A-B, |log_2_(FC)| > 0.25, Adjusted *P* < 0.05). It could be found that more proteins and metabolites were down-regulated in CC cases, indicating a general suppressive effect of normal biological and metabolic processes in children upon SARS-CoV-2 infection (Fig. 2A-B and Table S5-S6). However, up-regulated molecules exhibited stronger changes in expression, supporting that COVID-19-associated molecular alterations are not mild in children.

**Figure 2.**
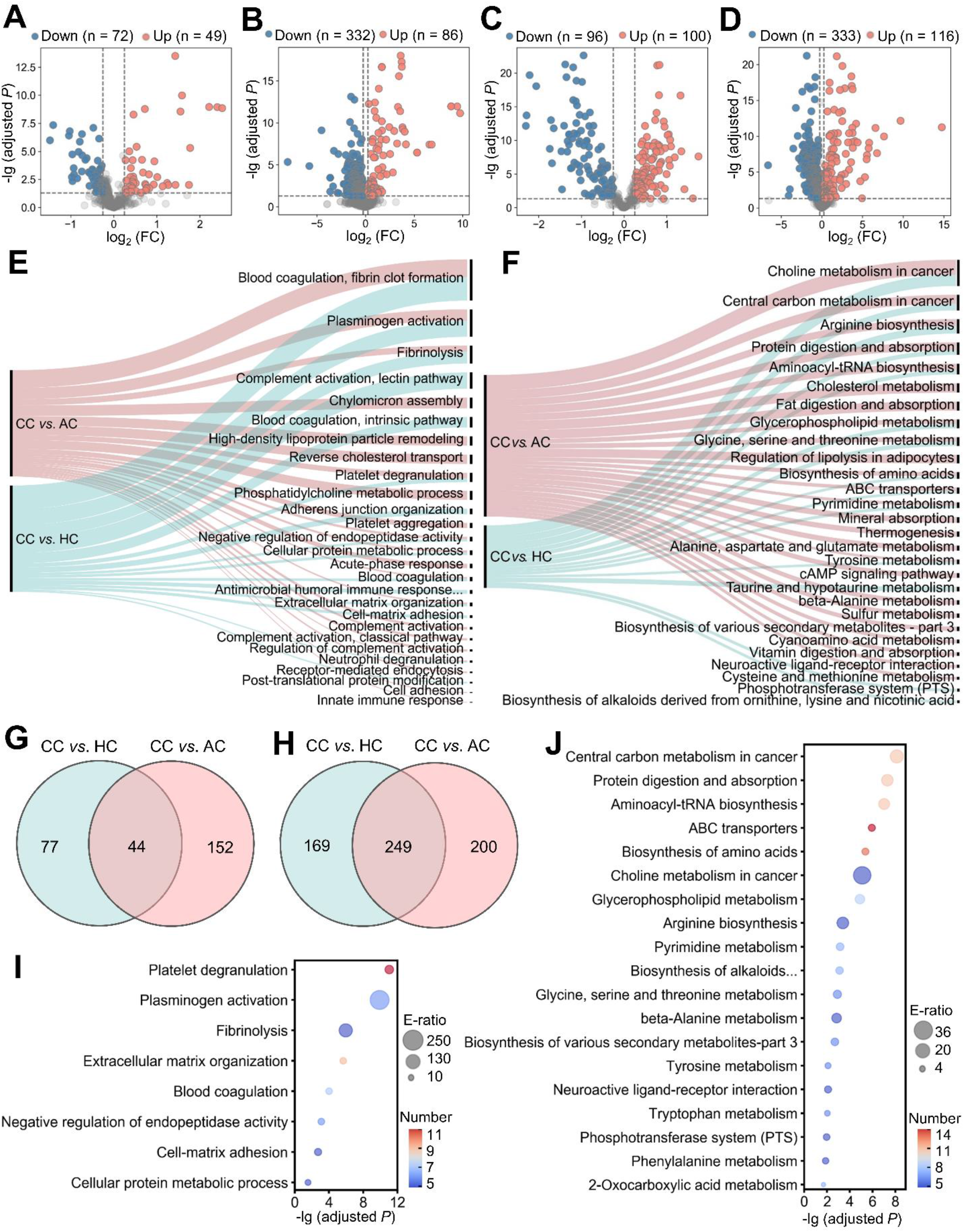
CC-specific proteomic and metabolomic alterations. **A**,**B** Volcano plots show the protein (A) and metabolite (B) alterations in CC against HC cases. **C**,**D** Volcano plots show the protein (C) and metabolite (D) alterations in CC against AC cases. Proteins and metabolites with log2 (FC) below 0.25 or beyond −0.25 with adjusted *P* lower than 0.05 were considered as significantly differential expression. **E** GO-based enrichment analysis for DEPs of CC against HC cases and CC against AC cases (Two-sided hypergeometric test, *m* ≥ 5, adjusted *P* < 10^−5^). **F** KEGG-based enrichment analysis for DEMs of CC against HC cases and CC against AC cases (Two-sided hypergeometric test, *m* ≥ 5, adjusted *P* < 10^−3^). **G**,**H** CC-specific DEPs (G) and DEMs (H) were identified by overlapping DEPs and DEMs of CC vs. HC and CC vs AC, respectively. **I** GO-based enrichment analysis of CC-specific DEPs shown in the term of biological processes (Two-sided hypergeometric test, *m* ≥ 5, adjusted *P* < 0.01). **J** KEGG-based enrichment analysis of CC-specific DEMs (Two-sided hypergeometric test, *m* ≥ 5, adjusted *P* < 0.01).

To further identify molecular alterations in CC against AC cases, the intrinsic differences between adults and children were eliminated by calculating normalized abundance values (NAVs) of proteins and metabolites in CC or AC cases against their counterparts in HC or HA samples. Again, 332 proteins and 783 metabolites simultaneously quantified in > 80% children and adults were reserved to ensure the data quality, and the PCA analysis demonstrated that CC and AC samples can be clearly distinguished either by the proteomic or metabolomic data (Fig. S3A-B). Using NAVs, we identified 196 DEPs and 449 DEMs in CC against HC cases (Fig. 2C-D, |log_2_(FC)| > 0.25, Adjusted *P* < 0.05, Table S7-S8). We found much more metabolites were down-regulated in CC cases, whereas up-regulated metabolites exhibited stronger changes in expression. This result not only support some strong molecular alterations in CC cases, but also suggested that different biological and metabolic processes are affected upon COVID-19 in CC against AC cases.

Next, we used the annotations of Gene Ontology (GO) biological processes and Kyoto Encyclopedia of Genes and Genomes (KEGG) pathways, and performed functional enrichment analyses for proteins and metabolites, respectively (Fig. 2E-F and Table S9-S10). We found that a considerable number of biological processes and metabolic pathways enriched in CC *vs*. AC and CC *vs*. HC cases were overlapped, such as platelet degranulation (GO:0002576), blood coagulation (GO:0007596), fibrinolysis (GO:0042730) and plasminogen activation (GO:0031639) in the proteomic level (Fig. 2E), and ABC transporters (KEGG ID: map02010), biosynthesis of amino acids (KEGG ID: map01230) and pyrimidine metabolism (KEGG ID: map00240) in the metabolic level (Fig. 2F). These enriched processes in CC and AC are consistent with the COVID-19-associated pathophysiological processes identified by other previous omics studies, and that further confirmed that COVID-19-associated molecular alterations occurred in CC cases, as did in AC cases.

We sought to examine the CC-specific molecules by overlapping DEPs and DEMs of CC vs. HC and CC vs AS, and identified 44 and 249 CC-specific DEPs and DEMs, respectively (Fig. 2G-H and Table S11-S12). We performed functional enrichment analyses for CC-specific DEPs (Fig. 2I) and DEMs (Fig. 2J and Table S9-S10), respectively. The biological processes and metabolic pathways based on these CC-specific molecules were mainly enriched in blood coagulation-related processes in the proteomic level, and anabolism-related processes involved in amino acid biosynthesis in the metabolic level (Fig. S3C-D), suggesting the importance of these physiological changes in children in response to COVID-19.

### Machine learning-based inference of CC-specific biomarker combinations

Although 44 and 249 CC-specific DEPs and DEMs were identified (Fig. 2G-H), different molecules were altered with distinct extents in CC cases. Identification of optimal biomarker combinations will not only be helpful for an accurate classification of different types of patients, but also for uncovering the pathogenesis of COVID-19 in children. Here, we developed a new computational pipeline named iBM, which consisted of three steps, including mutual DEPs or DEMs selection (MDS), candidate combination generation (CCG) to randomly select 10,000 biomarker combinations, and final combination prioritization (FCP) to get the protein or metabolite combination with a maximal accuracy and a minimal bias from the 5-fold cross-validation (Fig. 3A). The accuracy of a model was evaluated by calculating the total area under curve (AUC) value, and we also computed the total root mean squared error (RMSE) to measure the prediction bias. In the step of FCP, a widely used machine learning algorithm, penalized logistic regression (PLR)^25-27^, was used for model training and parameter optimization (Fig. 3A). The CC-specific biomarker combinations were separately determined for the proteomic and metabolic data.

**Figure 3.**
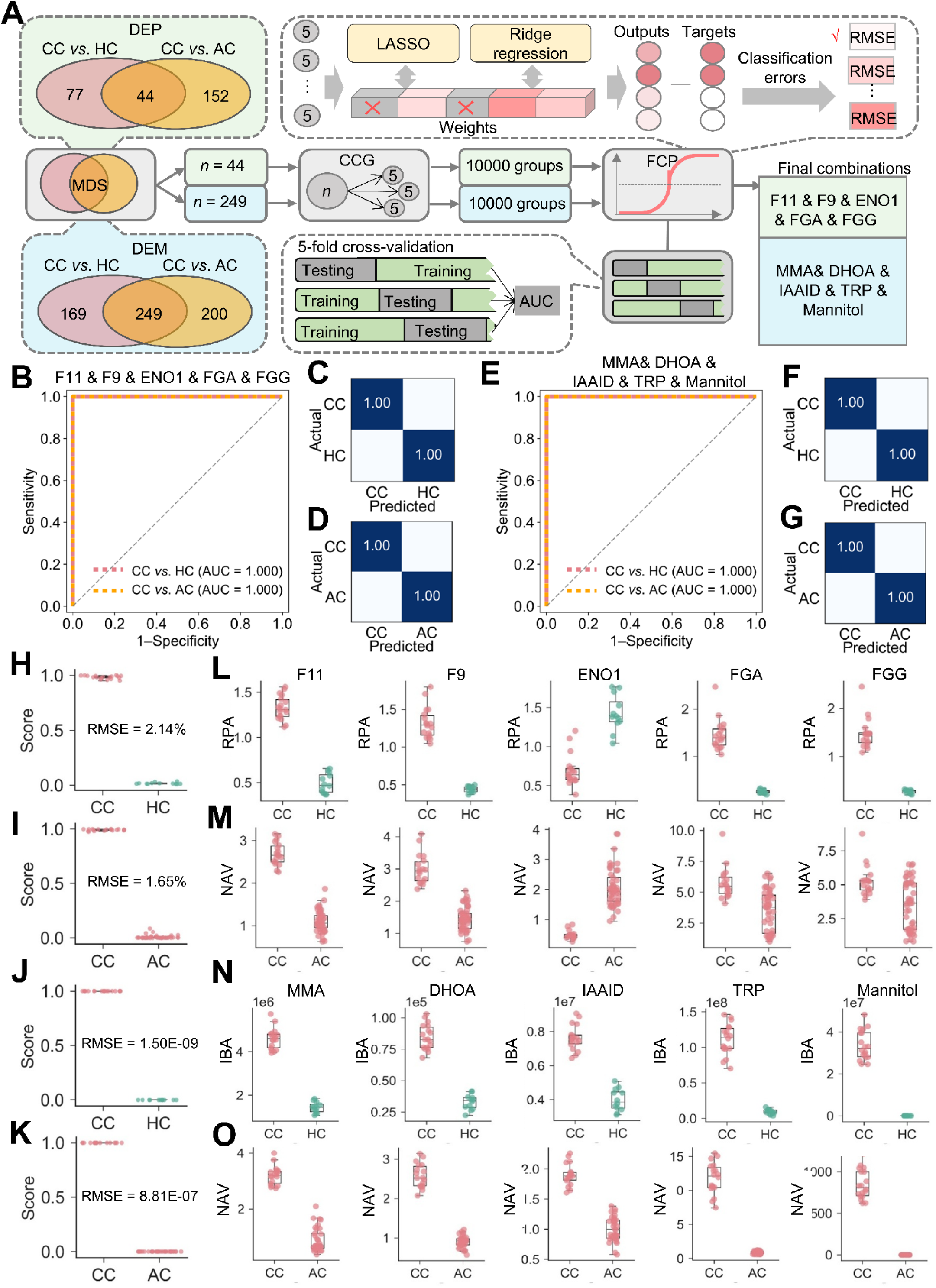
Inference of CC-specific biomarker combinations using a machine learning strategy. **A** The workflow of iBM, including MDS, CCG and FCP to prioritize CC-specific protein or metabolite biomarker combination with a maximal accuracy and a minimal bias from the 5-fold cross-validation. **B** From the 5-fold cross-validation, AUC values of protein combination for distinguishing the CC from HC cases and CC from AC cases were calculated, respectively. **C**,**D** The confusion matrix of the protein combination for distinguishing the CC from HC cases (C) and CC from AC cases (D). **E** From the 5-fold cross-validation, AUC values of metabolite combination for distinguishing the CC from HC cases and CC from AC cases were calculated, respectively. **F**,**G** The confusion matrices of the metabolite combination for distinguishing the CC from HC cases (F) and CC from AC cases (G). **H**,**I** The results of RMSE analyses of protein biomarker combinations for distinguishing the CC from HC cases (H) and CC from AC cases (I). **J**,**K** The results of RMSE analyses of metabolite biomarker combinations for distinguishing the CC from HC cases (J) and CC from AC cases (K). **L**,**M** The expression level of 5 protein biomarkers in CC against HC cases (L) and CC against AC cases (M). **N**,**O** The expression level of 5 metabolite biomarkers in CC against HC cases (N) and CC against AC cases (O). The center line within each box shows the median, and the top and bottom of each box represent the 75^th^ and 25^th^ percentile values, respectively. The upper and lower whiskers extend from the hinge to the largest and smallest value no further than 1.5 times the distance between the first and third quartiles, respectively.

From the results, there were 8098 protein combinations and 8376 metabolite combinations with a total AUC value of 1 (Table S13-S14), indicating that too many combinations could achieve a perfect accuracy on the existing data. However, a minimal RMSE value between predicted scores and observed values will ensure the robustness and reliability of the model on the new data. With total RMSE values of 1.83% and 7.01E-07, we prioritized two optimal biomarker combinations, containing 5 proteins coagulation factor XI and IX (F11 and F9), enolase (ENO1), fibrinogen alpha (FGA) and gamma (FGG) chains, and 5 metabolites methylmalonic acid (MMA), dihydroorotic acid (DHOA), indoleacetaldehyde (IAAID), tryptophan (TRP) and mannitol (Fig. 3A). Both of two biomarker combinations can perfectly distinguish CC group from AC and HC groups with AUC value of 1 (Fig. 3B-G). Moreover, the results of confusion matrices and RMSE analyses of these biomarker combinations also showed high accuracy for classifying different groups (Fig. 3C, D, F, G and H-O).

Also, we calculated the total AUC values and total RMSE values for individual proteins or metabolites. For the 5 proteins, the total AUC values ranged from 0.77 to 1, and the total RMSE values ranged from 6.57% to 36.13% (Fig. S4A-E, Table S15). For the 5 metabolites, all the total AUC values were 1, while the total RMSE values ranged from 10.97% to 28.72% (Fig. S4F-J, Table S15). Although individual molecules can reach a perfect accuracy on the current data, the combination of multiple molecules was undoubtedly important to reduce the prediction bias.

### The alterations of CC-specific molecules in plasma are linked with the mild symptoms of COVID-19 in children

The promising biomarkers for CC as well as other CC-specific molecules may also partially explain the differences in clinical symptoms between CC and AC cases. Of the protein biomarker combination, 4 proteins, including F11, F9, FGA and FGG were involved in the blood coagulation cascade, and all of them were higher expressed in CC cases than those in AC cases (Fig. 3L-M). F11 and F9 contribute the initiation of the thrombin generation during blood coagulation by proteolytic activation of a serial of coagulation factors^28^. FGA and FGG contribute to form the fibrin clot in response to explosive generation of thrombin mediated by coagulation factors^29^. Moreover, plasma serine protease inhibitor (SERPINA5) that negatively regulate the blood coagulation cascade^28^ was the most significantly upregulated one among all the CC-specific DEPs (Table S11). Thus, the remarkably alternations of coagulation-related proteins in CC plasma suggest that COVID-19-associated coagulation and the accompanying immune response/inflammation in CC are more active than those in AC.

On the other hand, a large number of CC-specific molecules implicated in antioxidant and/or anti-inflammatory processes were also significantly altered in CC. For example, ENO1, one of the components in the protein biomarker combination, was significantly downregulated in CC cases compared to that in AC cases (Fig. 3L-M). ENO1 is a key glycolytic enzyme in the last steps of the catabolic glycolytic pathway. Previous study showed that the suppression of ENO1 in pulmonary artery smooth muscle cells prevented the hypoxia-induced metabolic reprogramming from mitochondrial respiration to glycolysis^30^, the process of which enhance the oxidant stress and inflammation^31^. Moreover, for the components of the metabolite biomarker combination, DHOA that was significantly upregulated in CC cases (Fig. 3N-O), is involved in the pyrimidine metabolism and the secretion of DHOA can reduce the toxicity of glucose metabolism reprogramming in response to hypoxia^32^. TRP is metabolized to other indole compounds such as IAAID. The downstream products of TRP are key family of agonists to activate the aryl hydrocarbon receptor (AhR) that regulate the immunosuppression and restriction of inflammatory^33^. The remarkably elevated level of tryptophan and its downstream product indoleacetaldehyde suggested that AhR activity was more active in CC cases (Fig. 3N-O). Furthermore, mannitol has been found to be a hydroxyl radical scavenger that plays important roles in reducing inflammation^34^, and importantly, it was the most significantly upregulated one among all the CC-specific DEMs (Table S12). Besides, MMA is a dicarboxylic acid that is primarily a by-product of propionate metabolism. Previous study showed that the elevated circulating MMA level caused a significant upregulation of Sex-determining region Y box 4 (SOX4) expression and consequently elicited transcriptional reprogramming^35^, while the role of MMA in regulation of immune response and inflammation is unclear.

In addition to the molecules involved in the optimal biomarker combination, we also identified other molecules that contain the potentials to relieve the exacerbated inflammation. For instance, uric acid (UA), an end product of purine catabolism, is a major antioxidant in the blood and can be helpful for protection against free-radical oxidative damage^36^. The plasma level of uric acid in CC cases was 109-folds higher than that in AC cases (Table S12).

Together, our findings indicated that the molecules involved in anti-oxidant and anti-inflammation were significantly elevated in the circulating system in CC cases compared with AC cases. Consistent with it, the clinical data showed that the coagulation indicators, such as APTT, PT and D-dimer, the status of immune cell activation, such as the ratios of CD3+CD4+ and CD3+CD8+, and the levels of inflammatory factors, such as IFN-γ and IL-1β, in the CC cases were overall normal, albeit some results in certain cases fell outside the normal range (Table 1).

### Effects of regulating metabolism on coronaviral RNA replication and cytokine expressions *in vitro*

After identifying that CC-specific molecules may contribute to the mild COVID-19 symptom in children, it is intriguing to exploit the potential role of these metabolites on immune response and/or inflammation in the context of authentic viral infection. To this end, we used mouse hepatitis virus (MHV, strain A59)^37^, a well-known surrogate for SARS-CoV-2^38^, and tested the effects of MMA, DHOA and mannitol on the viral replication and cytokine expressions.

Rat lung epithelial L2 cells were treated with different metabolites at the concentration of 5 or 10 μM for 1 hr, and then infected with MHV at a multiplicity of infection (MOI) of 0.1, respectively. At 12 hrs post infection, total cellular RNAs were extracted and the viral RNA accumulation as well as the mRNA levels of IL-6, IL-1β, TNF-α, TNF-β and IL-10 were examined via using qRT-PCR. As expected, MHV infection resulted in significantly enhanced mRNA levels of cytokines (Fig. 4E-H). Moreover, all these metabolites can reduce the mRNA levels of cytokines in MHV-infected cells but not in mock cells, and the types of cytokines to change was different in response to treatment with distinct metabolites (Fig. 4E-G), suggesting that these metabolites regulate the expressions of cytokines via different pathways. Interestingly, while DHOA or TRP treatment showed no effect on MHV replication, we found that the RNA accumulation level of MHV was significantly reduced in the presence of MMA or mannitol (Fig. 4A, D). Together, these results provided experimental data to support that molecules involved in anti-inflammation were significantly elevated in CC.

**Figure 4.**
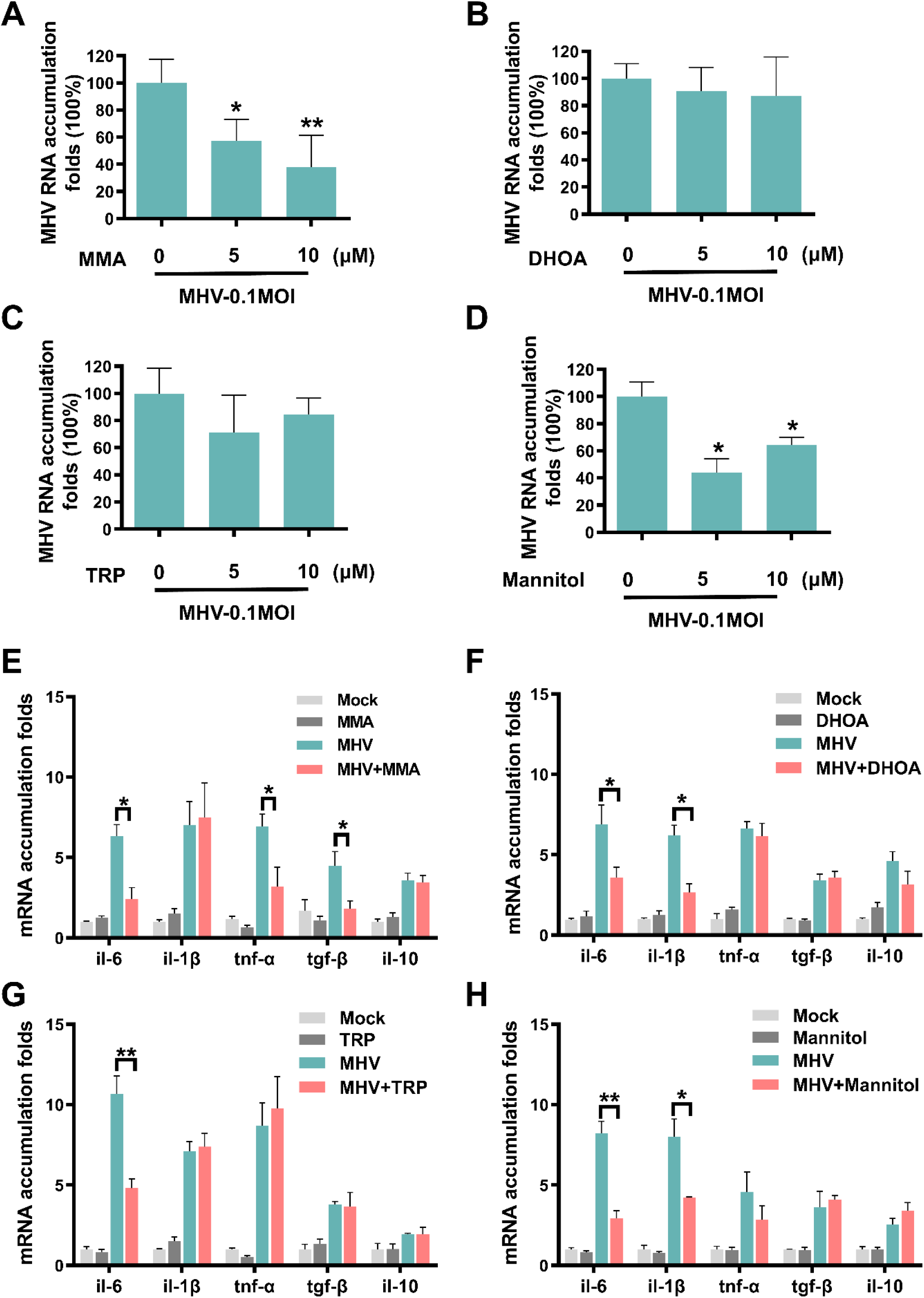
Effects of regulating metabolism on MHV RNA replication and cytokine expressions *in vitro*. **A-D** L2 cells were treated with MMA (A), DHOA (B), TRP (C) or mannitol (D) at the concentration of 5 or 10 μM, respectively, for 1 hr, and then infected with MHV at MOI=0.1, respectively. At 12 hr post infection, the total cellular RNAs were extracted. **E-H** The viral RNA accumulation and the mRNA levels of IL-6, IL-1β, TNF-α, TNF-β and IL-10 were determined via qRT-PCR. For measuring viral RNA replication, the level of MHV RNA in cells without treatment was defined as 100%. For measuring mRNA levels of cytokines, the mRNA level of each one of the tested cytokines in cells without infection was defined as 1-fold. The qRT-PCR and were measured by *t*-test (GraphPad Prism). \**P* < 0.05, \*\**P* < 0.01.

## Discussion

The infection rate of SARS-CoV-2 to children is similar to that of adults, but the disease of COVID-19 is very mild in most cases of CC. Therefore, better understanding the mechanisms underlying the milder COVID-19 symptom in children has particular importance to uncover the pathogenesis of this disease. For this purpose, we conducted omics study to profile plasma protein and metabolite alterations in CC cases, and identified CC-specific molecule alternations by comparing with the omics data of AC cases. Moreover, we prioritized two optimal biomarker combinations, each of them containing 5 protein biomarkers or 5 metabolite biomarkers, by using the machine learning-based pipeline. These biomarker combinations as well as the individual biomarker in the combinations can accurately distinguish CC group from AC and HC groups. Moreover, the alternations of these host molecules provide very valuable insight for the pathogenesis of COVID-19 in children. In addition, some of metabolite biomarkers were experimentally examined their roles in suppressing viral replication and/or modulating inflammation *in vitro*.

The identified alternations of plasma proteins and metabolites in this study as well as their enriched processes/pathways in CC are in line with the previous omics data with the plasma of AC cases^20, 21, 39-42^, indicating that COVID-19-associated molecular alterations occurred in children, as did in adults, in response to SARS-CoV-2 infection. However, we found that many molecules exhibited different changes in expression to different extents in CC cases compared to those in AC cases. Particularly, the plasma proteins involved in coagulation cascade were significantly higher expressed in CC cases compared with AC cases. Coagulation system plays important roles in immune responses against infections, and prevent damage to tissues and facilitate the repair of damaged areas^43^. While over-activation of coagulation cascade during the immune response to infection usually cause exacerbating production of pro-inflammatory cytokines, and coagulation-induced thrombin also exerts the activity to further augment inflammation^43^. Therefore, our findings suggest that COVID-19-associated coagulation and the accompanying immune response/inflammation in CC are strongly active. On the other hand, the levels of many negative regulators of inflammation and oxidation, such as TRP, IAAID, DHOA, mannitol and UA in CC cases were significantly upregulated compared with those in AC cases, indicating a feedback. Moreover, our findings showed that these metabolites not only can relieve the expressions of different pro-inflammatory factors, but also contain the unexpected activity to inhibit MHV replication in cells. These result further confirm that immune response in CC is strengthened and suggest that SARS-CoV-2 replication in CC is restricted by the enhanced levels of COVID-19-associated plasma molecules. Correspondingly, we speculated that the immune system of CC is in a relatively balanced state, in which its activation is stronger than that of AC and is sufficient to restrict SARS-CoV-2 infection and the collateral damages, and meanwhile, the molecules involved in anti-oxidant and anti-inflammation processes are also strongly activated in CC, thereby preventing the exacerbation of inflammation and the deterioration of disease (Fig. 5).

**Figure 5.**
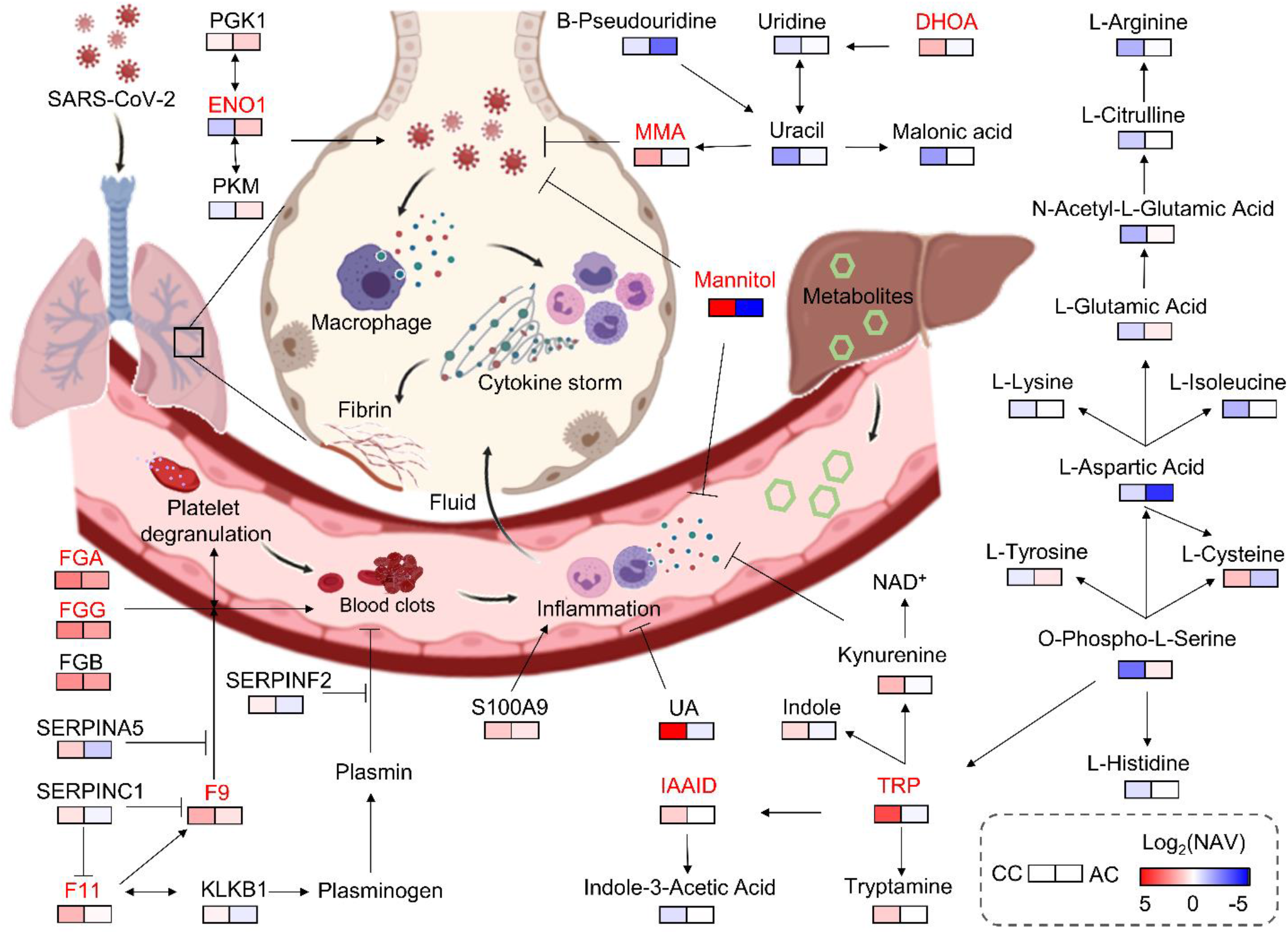
Key proteins and metabolites characterized in a working model associated with immune system of CC. In the working model, the plasma proteins involved in coagulation cascade were significantly higher expressed in CC cases compared with AC cases, suggesting COVID-19-associated coagulation and the accompanying immune response/inflammation in CC may be strongly active. On the other hand, the levels of many negative regulators of inflammation and oxidation, such as TRP, IAAID, DHOA, mannitol and UA in CC cases were significantly upregulated compared with those in AC cases, indicating a feedback. Moreover, metabolite Mannitol, MMA can enhance the IFN system and/or inhibit MHV replication with unknown mechanism in cells. As a whole, immune system of CC is in a relatively balanced state, in which its activation is stronger than that of AC and is sufficient to restrict SARS-CoV-2 infection and the collateral damages, and meanwhile, the molecules involved in anti-oxidant and anti-inflammation processes are also strongly activated in CC, thereby preventing the exacerbation of inflammation and the deterioration of disease.

Identification of the alternations of plasma molecules between CC and AC also provides promising therapeutic targets for COVID-19. In this study, we tested the effects of MMA, DHOA, TRP and mannitol on the expression levels of cytokines, as well as the viral replication in MHV-infected cells. Interestingly, the changes in the types of cytokine were different in response to distinct metabolite treatments, suggesting that the action to mode of these metabolites are dependent on different cellular signaling pathways. Moreover, we found that MMA or mannitol treatment can efficiently inhibit MHV replication. Mannitol is reported to be a hydroxyl radical scavenger that plays important roles in relieving inflammation^34^. A recent study showed that the SARS-CoV-2 infection in monocytes triggers mitochondrial ROS production, which induces stabilization of hypoxia-inducible factor-1α (HIF-1α) and consequently metabolism reprogramming that facilitates the viral replication and inhibits immune responses requires^44^. It is possible that the hydroxyl radical scavenging activity of mannitol relieves the cellular levels of ROS, and in turn suppresses MHV replication. For MMA, its exact role in regulation of immune response or inflammation is not clear. Previous study showed that MMA treatment in A549 cells triggered the induction of SOX4^35^, and interestingly, a recent study found that SOX4 can suppress hepatitis B virus replication via inhibiting hepatocyte nuclear factor 4α^45^. The detailed roles of the identified CC-specific metabolites such as MMA, DHOA and mannitol, require further investigation and the potential therapeutic targets should be further experimentally validated.

In summary, our findings provide highly valuable multi-omics data resource for the research community to better understand COVID-19-associated host responses, identify a serial of children-specific COVID-19 biomarker candidates, shed lights on the pathogenesis of COVID-19 in children, and provide promising potential therapeutic targets.

## Materials and Methods

### Ethics and Human Subjects

All work performed in this study was approved by the Guangzhou Women and Children’s Medical Center Ethics Committee and Written informed consent was waived by the Ethics Commission of the designated hospital for emerging infectious diseases. Diagnosis of SARS-CoV-2 infection was based on the New Coronavirus Pneumonia Prevention and Control Program (6th edition) published by the National Health Commission of China^22^. HC subjects were recruited at Guangzhou Women and Children’s Medical Center. The throat swabs and serological testing of H volunteers were negative for SARS-CoV-2. All blood samples were collected after fasting overnight and by added with ethylene diamine tetraacetic acid (EDTA) plus potassium (K^+^). All the blood samples were treated according to the biocontainment procedures of the processing of SARS-CoV-2-positive samples.

### Preparation of protein and peptide samples

Firstly, the cellular debris of serum sample was removed by centrifugation at 12,000 g at 4 °C for 10 min. Then, the supernatant was transferred to new centrifuge tubes. The top 12 high abundance proteins were removed by Pierce™ Top 12 Abundant Protein Depletion Spin Columns Kit (Thermo Fisher). Finally, the protein concentration was determined with BCA kit according to the manufacturer’s instructions.

For digestion, the protein solution was reduced with 5 mM dithiothreitol for 30 min at 56 °C and alkylated with 11 mM iodoacetamide for 15 min at room temperature in darkness. The protein sample was made buffer exchange by 8M UREA three times and then made buffer exchange by the label buffer three times. Finally, trypsin was added at 1:50 trypsin-to-protein mass ratio for the digestion overnight at 37?. Recover the peptide by centrifugation at 12,000 g at room temperature for 10 min and repeat the recovery step by H_2_O.

For TMT labeling, samples are divided to two groups according to the comparison design and processed according to the manufacturer’s protocol for TMT pro 16plex Label Reagent (Thermo Fisher) kit. Briefly, one unit of TMT pro reagent (defined as the amount of reagent required to label 100 μg of protein) were thawed and reconstituted in 10 μl ACN. The peptide mixtures were then incubated for 2 hrs at room temperature and pooled, desalted and dried by vacuum centrifugation. The labeling efficiency (calculated from the ratio of number of TMT labeled sites divided by number of all the potential labeling sites) had to pass the threshold of 95% before proceeding to the fractionation step.

The sample was then fractionated into fractions by high pH reverse-phase HPLC using Agilent 300Extend C18 column (5 μm particles, 4.6 mm ID, 250 mm length). Briefly, peptides were first separated with a gradient of 2% to 32% acetonitrile in 10 mM ammonium bicarbonate pH 10 over 60 min into 60 fractions^46^. Then, the peptides were combined into 6 fractions and dried by vacuum centrifuging.

### Extraction of hydrophilic and hydrophobic metabolites

To extract hydrophilic compounds, sample was thawed on ice, 6 volumes of ice-cold methanol was added to 1 volume of plasma, whirled the mixture for 3 min and centrifuge it at 12,000 rpm at 4°C for 5 min. Then collect the supernatant and leave in a refrigerator at - 20°C. After 30 min, centrifuge at 12000 r/min at 4°C for 3 min, and then collected the supernatant and subjected them to LC-MS/MS analysis.

To extract hydrophobic compounds, sample was thawed on ice, whirl around 10 s, and then centrifuge it with 3000 g at 4°C for 5 min. Take 50 μL of one sample and homogenized it with 1mL mixture (include methanol, MTBE and internal standard mixture). Whirled the mixture for 2 min. Then added 200 μL of water and whirled the mixture for 1 min, and centrifuged it with 12,000 g at 4°C for 10 min. Extracted 200 μL supernatant and concentrated it. Dissolved powder with 200 μL mobile phase B and subjected to LC-MS/MS analysis.

### LC-MS/MS-based proteomic analysis

LC-MS/MS data acquisition was carried out on a Exploris 480 mass spectrometer coupled with an Easy-nLC 1200 system (both Thermo Scientific)^40, 47^. Peptides were loaded onto a home-made reversed-phase analytical column (100 μm × 250 mm, 1.9 μm particle size, 120 Å pore size, Dr. Maish GmbH, Germany) and then separated. Mobile phase A (2% acetonitrile, 0.1% formic acid) and mobile phase B (90% acetonitrile, 0.1% formic acid) were used to establish a 60 min separation gradient (0 min – 7% B; 4 min – 11% B; 53 min – 32% B; 57 min – 80% B; 60 min – 80% B). A constant flow rate was set at 500 nL/min. For the analysis in data-dependent acquisition (DDA) mode, each scan cycle consisted of one full-scan mass spectrum (R = 60 K, AGC = 100%, max IT = 50 ms, scan range = 400–1200 m/z) followed by 25 MS/MS events (R = 45 K, AGC = 100%, max IT = Auto). High energy collision dissociation (HCD) collision energy was set to 35. Isolation window for precursor selection was set to 1.6 Da. Former target ion exclusion was set for 30 s.

### Protein database search

MS/MS raw data were analyzed with Proteome Discoverer (v2.4.1.15) using the Andromeda database search algorithm. The reference database contained 20,380 Swiss-Prot/reviewed human protein sequences downloaded from the UniProt database (https://www.uniprot.org/proteomes/UP000005640, on November 15, 2019), and reverse decoy sequences were generated. Then, spectra files were searched against the merged database using the following parameters: Type, TMT; Variable modifications, Oxidation (M), Acetyl (Protein N-term); Fixed modifications, Carbamidomethyl (C), TMTpro (peptide N-Terminus), TMTpro (K); Digestion, Trypsin (Full). The MS1 match tolerance was set as 10 parts per million (ppm); the MS2 tolerance was set as 0.02 Da. Search results were filtered with 1% false discovery rate (FDR) at both protein and peptide levels. Proteins denoted as decoy hits, or only identified by sites were removed, and the remaining proteins were used for further analysis.

### UPLC conditions of hydrophilic and hydrophobic compounds

The sample extracts of hydrophilic compounds were analyzed using an LC-ESI-MS/MS system (UPLC, Shim-pack UFLC SHIMADZU CBM A system, MS, QTRAP® 6500+ System). The samples were injected onto a Waters HSS T3 column (1.8 µm, 2.1 mm × 100 mm). Column temperature, flow rate and injection volume were set 40°C, 0.4 mL/min and 2 μL, respectively. Mobile phase was composed of water containing 0.1% formic acid (A) and acetonitrile containing 0.1% formic acid (B). The gradient program initiated from 5% B increased to 90% B in 11.0 min, and held for 1 min and then decreased 5% B for re-equilibrium.

The sample extracts of hydrophobic compounds were analyzed using an LC-ESI-MS/MS system (UPLC, Shim-pack UFLC SHIMADZU CBM A system, MS, QTRAP® 6500+ System). The samples were injected onto a Thermo Accucore™ C30 column (2.6 μm, 2.1 mm × 100 mm). Mobile phase was composed of acetonitrile/water (60/40, v/v) containing 0.1% formic acid, and 10 mmol/L ammonium formate (A) and acetonitrile/isopropanol (10/90, v/v) containing 0.1% formic acid and 10 mmol/L ammonium formate (B). The gradient program initiated A/B(80:20, V/V) at 0 min, 70:30 V/V at 2.0 min, 40:60 V/V at 4 min, 15:85 V/V at 9 min, 10:90 V/V at 14 min, 5:95 V/V at 15.5 min, 5:95 V/V at 17.3 min, 80:20 V/V at 17.3 min, 80:20 V/V at 20 min. The flow rate, column temperature and injection volume were set 0.35 ml/min, 45°C and 2 μL, respectively. The effluent was alternatively connected to an ESI-triple quadrupole-linear ion trap (QTRAP)-MS.

### ESI-Q TRAP-MS/MS of hydrophilic and hydrophobic compounds

LIT and triple quadrupole (QQQ) scans were acquired on a triple quadrupole-linear ion trap mass spectrometer (QTRAP), QTRAP® LC-MS/MS System, equipped with an ESI Turbo Ion-Spray interface, operating in positive and negative ion modes and controlled by Analyst 1.6.3 software (Sciex). The ESI source operation parameters were as follows: ion source, turbo spray; source temperature 550°C; ion spray voltage (IS) 5500 V in positive ion mode (or −4500 V in negative ion mode); ion source gas I (GSI), gas II (GSII), curtain gas (CUR) were set at 45, 55, and 35 psi, respectively; the collision gas (CAD) was medium. Instrument tuning and mass calibration were performed with 10 and 100 μM polypropylene glycol solutions in QQQ and LIT modes, respectively. QQQ scans were acquired as MRM experiments with collision gas (nitrogen) set to 5 psi. Declustering potential (DP) and collision energy (CE) for individual MRM transitions was done with further DP and CE optimization. A specific set of MRM transitions were monitored for each period according to the metabolites within this period. Each sample analysis was conducted by both positive and negative ion modes, and the MRM transitions were listed in Table S16.

### Plasma metabolites and lipids data analysis

The mass spectrum data were processed by Software Analyst 1.6.3. The repeatability of metabolite extraction and detection can be judged by total ion current (TIC) and multi peak MRM. Based on home-made MWDB (metadata database) and other databases, qualitative analysis of information and secondary general data was carried out according to retention time (RT) and mass-to-charge ratio. Metabolite structure analysis referred to some existing mass spectrometry public databases, mainly including massbank (http://www.massbank.jp/), knapsack (http://kanaya.naist.jp/knapsack/), HMDB (http://www.hmdb.ca/), and Metlin (http://metlin.scripps.edu/index.php). The metabolite identification was conducted by reference standards in our home-made database and public databases (Table S4).

For the quality control (QC) of metabolomic analysis, we pipette 10 μL of each sample to pool a QC sample. When running sample sets on column, one QC sample was injected after 10 samples in the sequence. Metabolite quantification was accomplished by using multiple reaction monitoring (MRM) of triple quadrupole mass spectrometry. Opened the mass spectrum file under the sample machine with multiquant software to integrate and calibrate the chromatographic peaks. The peak area of each chromatographic peak represented the relative content of the corresponding substance. Finally, exported all the integral data of chromatographic peak area to save, and used the self-built software package to remove the positive and negative ions of metabolites. We calculated CV values of the metabolites in QC samples, and removed the metabolites whose CV values were larger than 0.5. When the metabolites were detected in both positive and negative ionization modes, we removed the metabolites with larger CVs in either positive or negative mode.

### Cell culture, virus and infection

L2 cell line was kindly provided by Prof. Chen (Wuhan University, China) and maintained in Dulbecco’s modified Eagle’s medium (DMEM) (Gibco) supplemented with 10% fetal bovine serum (FBS) (Gibco), 100 U/ml penicillin and 100 μg/ml streptomycin at 37°C in a humidified atmosphere with 5% CO_2_. MHV strain A59 was kindly provided by Prof. Chen (Wuhan University, China).

MMA (STBF5304V), DHOA (SLCD3296) and mannitol (WXBD1141V) were commercially purchased from Sigma-Aldrich. TRP (10211562) was commercially purchased from Alfa Aesar. For detection of the effects of MMA, DHOA, TRP and mannitol upon MHV infection, each one of the tested compounds at the concentration of 5 or 10 μM was added to L2 cells. After 1 hr incubation, L2 cells were infected with MHV at MOI=0.1. At 12 hr post infection, the infected L2 cells were collected and total cellular RNAs were extracted. The viral RNA accumulation and the mRNA levels of IL-6, IL-1β, TNF-α, TNF-β and IL-10 were determined via qRT-PCR. For measuring viral RNA replication, the level of MHV RNA in cells without treatment was defined as 100%. For measuring mRNA levels of cytokines, the mRNA level of each one of the tested cytokines in cells without infection was defined as 1-fold.

### Data normalization and imputation

For each batch of the plasma proteomic data, the IBA of a protein in one sample was first normalized using its corresponding expression in the control of the same batch to calculate the RPA, which eliminated the batch effect prior to the comparative analysis of CC and HC samples.

To identify molecular alterations exclusively in CC but not HC samples, the mean RPA value of a protein *i* in HC or HA samples was first calculated as below:

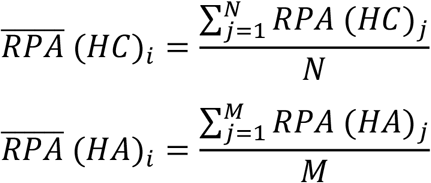

Where *N* and *M* denoted the numbers of HC and HA samples, respectively. Then, the NAV of the protein *i* in each CC or AC sample was calculated as below:

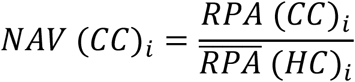

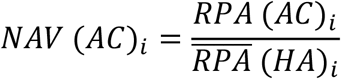

Analogously, the NAVs of all metabolites in each CC or AC samples were also computed. To ensure the data quality for identification of potential DEPs or DEMs, we only reserved proteins or metabolites quantified in > 80% samples (> 24 samples for CC *vs*. HC analysis, > 68 samples for the proteomic analysis of CC *vs*. AC, and > 59 samples for the metabolomic analysis of CC *vs*. AC). Using NI, the missing values were imputed with values representing a normal distribution around the detection limit of the mass spectrometer. For each sample, the mean and standard deviation (S.D.) of the distribution of the raw protein or metabolite intensities were calculated. Then a new distribution with a downshift of 1.8 S.D. and a width of 0.3 S.D. was automatically modeled. The total data set was imputed before statistical analysis. After imputation, the mean *μ* and S.D *σ* were counted for each protein or metabolite in CC and HC samples, respectively, and CV was calculated as below:

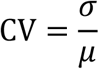

Before model training, the proteomic or metabolomic data of each sample was further normalized using the z-score transformation, one of the mostly used normalization methods^43^. For each sample, the median expression value *m* and S.D. *δ* were first calculated for the proteomic or metabolomic data. For a protein or metabolite *i* with the abundance of *NAV*_*i*_, its normalized *z*-score was calculated as below:

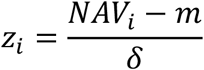

After transformation, the *z*-scores of proteins or metabolites followed a logarithmic normal distribution (log_2_) centered at zero.

The proteomic and metabolomic data normalization and imputation were conducted using Perseus 1.6.14^24^. To test whether different types of patients could be distinguished, PCA was performed using Scikit-learn 0.22.1 (https://scikit-learn.org/stable/), a useful toolkit for data mining and analysis. The Pearson correlation analysis was performed by an R packge, corrplot (https://cran.r-project.org/web/packages/corrplot/index.html).

### Statistical analysis of the quantitative omic data

Using RPA values of the proteomic data and IBA values of the metabolomic data, we identified potential DEPs and DEMs that were significantly altered in CC cases against HC samples. Then, using NAVs of the omic data, we further identified potential DEPs and DEMs that were significantly altered in CC cases against AC cases. The FC value was calculated based on the mean of the same patient group for each pair of groups, and proteins or metabolites with |log_2_(FC)| > 0.25 were reserved. The statistical significance was calculated for reserved proteins and metabolites, using the unpaired two-sided Welch’s t-test and adjusted *P* values were calculated using Benjamini & Hochberg correction (Adjusted *P* < 0.05). The statistical analyses were conducted using the ttest_ind function in scipy.stats.

### The enrichment analysis

The two-sided hypergeometric test was adopted for the GO- or KEGG-based enrichment analysis of the DEPs or DEMs. Here, we defined:

*N* = number of human proteins or metabolites annotated by at least one term

*n* = number of human proteins or metabolites annotated by term *t*

*M* = number of the DEPs or DEMs annotated by at least one term

*m* = number of the DEPs or DEMs annotated by term *t*

Then, the E-ratio was calculated, and the *P* value was computed with the hypergeometric distribution as below:

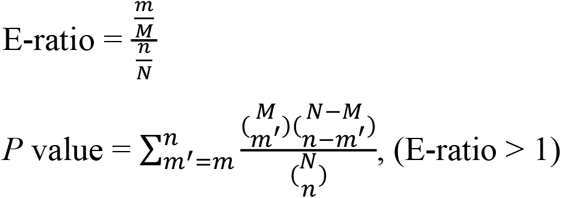

In this study, adjusted *P* values were calculated using Benjamini & Hochberg correction and only statistically over-represented GO terms for the proteomic data and KEGG pathways for the metabolomic data were considered. GO annotation files (released on 03 January 2020) were downloaded from the Gene Ontology Consortium Web site (http://www.geneontology.org/), and in total we obtained 19,288 human proteins annotated with at least one GO biological process term. KEGG annotation files (released on 4 September 2020) were downloaded from the ftp server of KEGG (ftp://ftp.bioinformatics.jp/), which contained 6,182 metabolites annotated with at least one KEGG pathway term.

### Performance evaluation

To evaluate the accuracy of iBM, the numbers of true positive (*TP*), true negative (*TN*), false positive (*FP*) and false negative (*FN*) hits were counted. Then, we calculated two measurements, including sensitivity (*Sn*), specificity (*Sp*) as below:

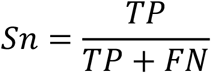

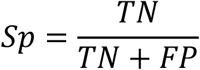

The 5-fold cross-validation was performed, while *Sn* and *Sp* values were calculated, respectively. The receiver operating characteristic (ROC) curve was illustrated and AUC value was calculated based on *Sn* and 1-*Sp* scores.

To estimate the prediction bias of a model, the root mean squared error (RMSE), an important measure of the residuals between predicted values and observed values, was calculated as below:

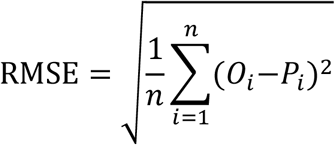

Where *n* represented the number of plasma samples in the data set. *P*_*i*_ denoted the predicted probability value ranged from 0 to 1, while *O*_*i*_ was equal to 0 for non-COVID-19 cases and 1 for COVID-19 cases, respectively.

### Inference of optimal biomarker combinations

We separately identified potentially CC-specific biomarker combinations with minimal RMSE for the proteomic and metabolomic data, by developing a three-step pipeline named iBM that included mutual DEPs or DEMs selection (MDS), candidate combination generation (CCG), and final combination prioritization (FCP).

In the step of MDS, mutually identified DEPs or DEMs of CC patients against HC cases and CC cases against AC cases were reserved as a candidate pool. Then, CCG was adopted to select different sets of biomarker combinations with ≤ 5 proteins or metabolites. This number was much smaller than the sample size and could efficiently avoid over-fitting. From the pool, 10,000 candidate biomarker combinations were randomly generated for the proteomic and metabolomic data, respectively. The initial weight value of each protein or metabolite was set to 1.

In the step of FCP, the 5-fold cross-validation was conducted for model training. For each candidate combination, we randomly generated a training data set and a testing data set with a ratio of approximately 4:1. The testing data set was only used to test the performance but not for training, and the final total AUC value was calculated as below:

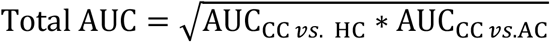

The least absolute shrinkage and selection operator (LASSO, L1 regularization) penalty and the ridge regression (L2 regularization) penalty in PLR^25-27^, were iteratively used to optimize the weight values of the 5 proteins or metabolites. To simplify the composition of a combination, one or multiple protein or metabolite was randomly dropped if the total AUC value of the 5-fold cross-validation was increased. Such a procedure was repeatedly performed until the AUC value was not increased any longer. All biomarker combinations with total AUC equal to 1 were reserved for the proteomic and metabolomic data, respectively (Table S11-S12). The total RMSE value of all samples was calculated for each combination, and the final result was determined based on the minimal total RMSE value.

The PLR algorithm was implemented in Python 3.7 with Scikit-learn 0.22.1. The source code of iBM is available at: https://github.com/Ning-310/iBM.

### Construction of a working model associated with immune system of CC

We constructed a working model of CC-specific immune response around 5 CC-specific protein biomarkers and 5 CC-specific metabolite biomarkers. Based on the pathway annotations in KEGG, working model included the 13 CC-specific DEPs and 25 CC-specific DEMs mainly involved in 6 KEGG pathways, including platelet activation, complement and coagulation cascades, glycolysis/gluconeogenesis, pyrimidine metabolism, biosynthesis of amino acids and tryptophan metabolism.

## Data Availability

The mass spectrometry proteomics data have been deposited to the ProteomeXchange Consortium (http://proteomecentral.proteomexchange.org) via the iProX partner repository with the project ID: IPX0002173000, IPX0002243000 and IPX0002673000.

## General

We thank many staff members at Guangzhou Women and Children’s Medical Center their contributions and Prof. Yu Chen (Wuhan university) for his kind assistance in this study. We also thank SpecAlly Life Technology Co., Ltd. and Wuhan Metware Biotechnology Co., Ltd. for their contributions and assistance in this study.

## Funding

This work was supported by the Strategic Priority Research Program of CAS (XDB29010300 to X.Z.), the National Science and Technology Major Project (2018ZX10101004 to X.Z.), National Natural Science Foundation of China (81873964 to Y.Q., 31930021, 31970633 and 34671360 to Y.X., and 31670161 to X.Z.), Grant from the CAS Youth Innovation Promotion Association (2020332 to Y.Q.), the program for HUST Academic Frontier Youth Team (Y.X.).

## Author contributions

C.W., W.N. and X.L. performed experiments with the help of S.G., D.W., M.H., F.Y., C.F., Y.G., Y.R. and R.Y.; Y.X., W.N., and Y.Q. analyzed the data with the help of C.W. and X.L.; Y.Q., W.N., Y.X., C.W. and X.Z. wrote the manuscript; X.Z., Y.Xu., Y.X. and Y.Q. designed and supervised the overall study.

## Competing interests

The authors declare no conflicts of interest.

## Data and materials availability

The mass spectrometry proteomics data have been deposited to the ProteomeXchange Consortium (http://proteomecentral.proteomexchange.org) via the iProX partner repository with the dataset identifier (https://www.iprox.org//page/project.html?id=IPX0002673000).

## Notes

### Competing Interest Statement

The authors have declared no competing interest.

### Clinical Trial

21600

### Author Declarations

All work performed in this study was approved by the Guangzhou Women and Children's Medical Center Ethics Committee and Written informed consent was waived by the Ethics Commission of the designated hospital for emerging infectious diseases.

## References

1 Y. Dong, X. Mo, Y. Hu, X. Qi, F. Jiang, Z. Jiang, and S. Tong, Pediatrics. 145, (2020)

2 K. Yuki, M. Fujiogi, and S. Koutsogiannaki, Clin Immunol. 215, 108427 (2020)

3 J. F. Ludvigsson, Acta Paediatr. 109, 1088–1095 (2020)

4 T. C. Jones, B. Mühlemann, T. Veith, G. Biele, M. Zuchowski, J. Hoffmann, A. Stein, A. Edelmann, V. M. Corman, and C. Drosten, medRxiv. (2020)

5 L. Su, X. Ma, H. Yu, Z. Zhang, P. Bian, Y. Han, J. Sun, Y. Liu, C. Yang, J. Geng, Z. Zhang, and Z. Gai, Emerging Microbes & Infections. 9, 707–713 (2020)

6 Y. Dong, X. Mo, Y. Hu, X. Qi, F. Jiang, Z. Jiang, and S. Tong, Pediatrics. 145, (2020)

7 Y. Xu, X. Li, B. Zhu, H. Liang, C. Fang, Y. Gong, Q. Guo, X. Sun, D. Zhao, J. Shen, H. Zhang, H. Liu, H. Xia, J. Tang, K. Zhang, and S. Gong, Nat Med. 26, 502–505 (2020)

8 H. Qiu, J. Wu, L. Hong, Y. Luo, Q. Song, and D. Chen, Lancet Infect Dis. 20, 689–696 (2020)

9 N. Parri, M. Lenge, and D. Buonsenso, N Engl J Med. 383, 187–190 (2020)

10 Q. Wu, Y. Xing, L. Shi, W. Li, Y. Gao, S. Pan, Y. Wang, W. Wang, and Q. Xing, Pediatrics. 146, (2020)

11 H. Ma, J. Hu, J. Tian, X. Zhou, H. Li, M. T. Laws, L. D. Wesemann, B. Zhu, W. Chen, R. Ramos, J. Xia, and J. Shao, BMC Med. 18, 123 (2020)

12 E. Whittaker, A. Bamford, J. Kenny, M. Kaforou, C. E. Jones, P. Shah, P. Ramnarayan, A. Fraisse, O. Miller, P. Davies, F. Kucera, J. Brierley, M. McDougall, M. Carter, A. Tremoulet, C. Shimizu, J. Herberg, J. C. Burns, H. Lyall, and M. Levin, JAMA. 324, 259–269 (2020)

13 E. W. Cheung, P. Zachariah, M. Gorelik, A. Boneparth, S. G. Kernie, J. S. Orange, and J. D. Milner, JAMA. 324, 294–296 (2020)

14 C. R. Consiglio, N. Cotugno, F. Sardh, C. Pou, D. Amodio, L. Rodriguez, Z. Tan, S. Zicari, A. Ruggiero, G. R. Pascucci, V. Santilli, T. Campbell, Y. Bryceson, D. Eriksson, J. Wang, Marchesi, T. Lakshmikanth, A. Campana, A. Villani, P. Rossi, N. Landegren, P. Palma, and P. Brodin, Cell. 183, 968–981 e7 (2020)

15 P. Brodin, Acta Paediatr. 109, 1082–1083 (2020)

16 A. K. Simon, G. A. Hollander, and A. McMichael, Proc Biol Sci. 282, 20143085 (2015)

17 A. Olin, E. Henckel, Y. Chen, T. Lakshmikanth, C. Pou, J. Mikes, A. Gustafsson, A. K. Bernhardsson, C. Zhang, K. Bohlin, and P. Brodin, Cell. 174, 1277–1292 e14 (2018)

18 S. Nickbakhsh, C. Mair, L. Matthews, R. Reeve, P. C. D. Johnson, F. Thorburn, B. von Wissmann, A. Reynolds, J. McMenamin, R. N. Gunson, and P. R. Murcia, Proc Natl Acad Sci U S A. 116, 27142–50 (2019)

19 S. Bunyavanich, A. Do, and A. Vicencio, JAMA. 323, 2427–2429 (2020)

20 D. Wu, T. Shu, X. B. Yang, J. X. Song, M. L. Zhang, C. Y. Yao, W. Liu, M. H. Huang, Y. Yu, Q. Y. Yang, T. J. Zhu, J. Q. Xu, J. F. Mu, Y. X. Wang, H. Wang, T. Tang, Y. J. Ren, Y. R. Wu, S. H. Lin, Y. Qiu, D. Y. Zhang, Y. Shang, and X. Zhou, National Science Review. 7, 1157–1168 (2020)

21 T. Shu, W. Ning, D. Wu, J. Xu, Q. Han, M. Huang, X. Zou, Q. Yang, Y. Yuan, Y. Bie, S. Pan, J. Mu, Y. Han, X. Yang, H. Zhou, R. Li, Y. Ren, X. Chen, S. Yao, Y. Qiu, D. Y. Zhang, Y. Xue, Y. Shang, and X. Zhou, Immunity. 53, 1108–1122 e5 (2020)

22 T. National Health Commission, National Health Commission of the People’s Republic of China. (2020)

23 S. Tyanova, T. Temu, and J. Cox, Nat Protoc. 11, 2301–2319 (2016)

24 S. Tyanova, T. Temu, P. Sinitcyn, A. Carlson, M. Y. Hein, T. Geiger, M. Mann, and J. Cox, Nature Methods. 13, 731–740 (2016)

25 W. S. Ning, S. J. Lei, J. J. Yang, Y. K. Cao, P. R. Jiang, Q. Q. Yang, J. Zhang, X. B. Wang, F. H. Chen, Z. Geng, L. Xiong, H. M. Zhou, Y. P. Guo, Y. L. Zeng, H. S. Shi, L. Wang, Y. Xue, and Z. Wang, Nature Biomedical Engineering. (2020)

26 W. Ning, P. Jiang, Y. Guo, C. Wang, X. Tan, W. Zhang, D. Peng, and Y. Xue, Brief Bioinform. (2020)

27 W. S. Ning, H. D. Xu, P. R. Jiang, H. Cheng, W. K. Deng, Y. P. Guo, and Y. Xue, Genomics Proteomics & Bioinformatics. 18, 194–207 (2020)

28 B. M. Mohammed, A. Matafonov, I. Ivanov, M. F. Sun, Q. Cheng, S. K. Dickeson, C. Li, D. Sun, I. M. Verhamme, J. Emsley, and D. Gailani, Thromb Res. 161, 94–105 (2018)

29 D. Davalos, and K. Akassoglou, Semin Immunopathol. 34, 43–62 (2012)

30 J. Dai, Q. Zhou, J. Chen, M. L. Rexius-Hall, J. Rehman, and G. Zhou, Nat Commun. 9, 3850 (2018)

31 J. Van den Bossche, L. A. O’Neill, and D. Menon, Trends Immunol. 38, 395–406 (2017)

32 Y. Wang, C. Bai, Y. Ruan, M. Liu, Q. Chu, L. Qiu, C. Yang, and B. Li, Nat Commun. 10, 201 (2019)

33 J. E. Cheong, and L. Sun, Trends Pharmacol Sci. 39, 307–325 (2018)

34 P. André, and F. Villain, Int J Cosmet Sci. 39, 355–360 (2017)

35 A. P. Gomes, D. Ilter, V. Low, J. E. Endress, J. Fernández -García, A. Rosenzweig, T. Schild, D. Broekaert, A. Ahmed, M. Planque, I. Elia, J. Han, C. Kinzig, E. Mullarky, A. P. Mutvei, J. Asara, R. de Cabo, L. C. Cantley, N. Dephoure, S. M. Fendt, and J. Blenis, Nature. 585, 283–287 (2020)

36 T. Mikami, and M. Sorimachi, Physiol Res. 66, 1001–1007 (2017)

37 L. Cao, Y. Ji, L. Zeng, Q. Liu, Z. Zhang, S. Guo, X. Guo, Y. Tong, X. Zhao, C. M. Li, Y. Chen, and D. Guo, PLoS Pathog. 15, e1008079 (2019)

38 R. W. Körner, M. Majjouti, M. A. A. Alcazar, and E. Mahabir, Viruses. 12, (2020)

39 B. Shen, X. Yi, Y. Sun, X. Bi, J. Du, C. Zhang, S. Quan, F. Zhang, R. Sun, L. Qian, W. Ge, W. Liu, S. Liang, H. Chen, Y. Zhang, J. Li, J. Xu, Z. He, B. Chen, J. Wang, H. Yan, Y. Zheng, D. Wang, J. Zhu, Z. Kong, Z. Kang, X. Liang, X. Ding, G. Ruan, N. Xiang, X. Cai, H. Gao, L. Li, S. Li, Q. Xiao, T. Lu, Y. Zhu, H. Liu, H. Chen, and T. Guo, Cell. 182, 59–72 e15 (2020)

40 D. E. Gordon, G. M. Jang, M. Bouhaddou, J. Xu, K. Obernier, K. M. White, M. J. O’Meara, V. V. Rezelj, J. Z. Guo, D. L. Swaney, T. A. Tummino, R. Huettenhain, R. M. Kaake,L. Richards, B. Tutuncuoglu, H. Foussard, J. Batra, K. Haas, M. Modak, M. Kim, P. Haas, B.J. Polacco, H. Braberg, J. M. Fabius, M. Eckhardt, M. Soucheray, M. J. Bennett, M. Cakir, M. J. McGregor, Q. Li, B. Meyer, F. Roesch, T. Vallet, A. Mac Kain, L. Miorin, E. Moreno, Z. Z. C. Naing, Y. Zhou, S. Peng, Y. Shi, Z. Zhang, W. Shen, I. T. Kirby, J. E. Melnyk, J. S. Chorba, K. Lou, S. A. Dai, I. Barrio-Hernandez, D. Memon, C. Hernandez-Armenta, J. Lyu, C. J. P. Mathy,T. Perica, K. B. Pilla, S. J. Ganesan, D. J. Saltzberg, R. Rakesh, X. Liu, S. B. Rosenthal, L. Calviello, S. Venkataramanan, J. Liboy-Lugo, Y. Lin, X. P. Huang, Y. Liu, S. A. Wankowicz, M. Bohn, M. Safari, F. S. Ugur, C. Koh, N. S. Savar, Q. D. Tran, D. Shengjuler, S. J. Fletcher, M. C. O’Neal, Y. Cai, J. C. J. Chang, D. J. Broadhurst, S. Klippsten, P. P. Sharp, N. A. Wenzell, D. Kuzuoglu, H. Y. Wang, R. Trenker, J. M. Young, D. A. Cavero, J. Hiatt, T. L. Roth, U. Rathore,Subramanian, J. Noack, M. Hubert, R. M. Stroud, A. D. Frankel, O. S. Rosenberg, K. A. Verba, D. A. Agard, M. Ott, M. Emerman, N. Jura, M. von Zastrow, E. Verdin, A. Ashworth, O. Schwartz, C. d’Enfert, S. Mukherjee, M. Jacobson, H. S. Malik, D. G. Fujimori, T. Ideker, C. S. Craik, S. N. Floor, J. S. Fraser, J. D. Gross, A. Sali, B. L. Roth, D. Ruggero, J. Taunton, T. Kortemme, P. Beltrao, M. Vignuzzi, A. García -Sastre, K. M. Shokat, B. K. Shoichet, and N. J. Krogan, Nature. (2020)

41 D. Bojkova, K. Klann, B. Koch, M. Widera, D. Krause, S. Ciesek, J. Cinatl, and C. Münch, Nature. (2020)

42 D. Kim, J. Y. Lee, J. S. Yang, J. W. Kim, V. N. Kim, and H. Chang, Cell. 181, 914–921 e10 (2020)

43 B. Arneth, Inflamm Res. 68, 117–123 (2019)

44 A. C. Codo, G. G. Davanzo, L. B. Monteiro, G. F. de Souza, S. P. Muraro, J. V. Virgilio-da-Silva, J. S. Prodonoff, V. C. Carregari, C. A. O. de Biagi Junior, F. Crunfli, J. L. Jimenez Restrepo, P. H. Vendramini, G. Reis-de-Oliveira, K. Bispo Dos Santos, D. A. Toledo-Teixeira, P.L. Parise, M. C. Martini, R. E. Marques, H. R. Carmo, A. Borin, L. D. Coimbra, V. O. Boldrini, N. S. Brunetti, A. S. Vieira, E. Mansour, R. G. Ulaf, A. F. Bernardes, T. A. Nunes, L. C. Ribeiro, C. Palma, M. V. Agrela, M. L. Moretti, A. C. Sposito, F. B. Pereira, L. A. Velloso, M. A. R. Vinolo, A. Damasio, J. L. Proença -Módena, R. F. Carvalho, M. A. Mori, D. Martins-de-Souza, H. Nakaya, A. S. Farias, and P. M. Moraes-Vieira, Cell Metab. 32, 498–499 (2020)

45 S. Shi, M. Liu, J. Xi, H. Liu, G. Guan, C. Shen, Z. Guo, T. Zhang, Q. Xu, D. Kudereti, X. Chen, J. Wang, and F. Lu, Antiviral Research. 176, 104745 (2020)

46 T. S. Batth, C. Francavilla, and J. V. Olsen, J Proteome Res. 13, 6176–86 (2014)

47 M. Miao, F. Yu, D. Wang, Y. Tong, L. Yang, J. Xu, Y. Qiu, X. Zhou, and X. Zhao, Virol Sin. 34, 549–562 (2019)

