## Supplemental Figures for "Multi-Omic Profiling of Plasma Identify Biomarkers and Pathogenesis of COVID-19 in Children"

**This PDF file includes:**

Figs. S1 to S4

**Other Supplementary Materials for this manuscript include the following:**

Datasheet Table S1 to S16

### Figure S1

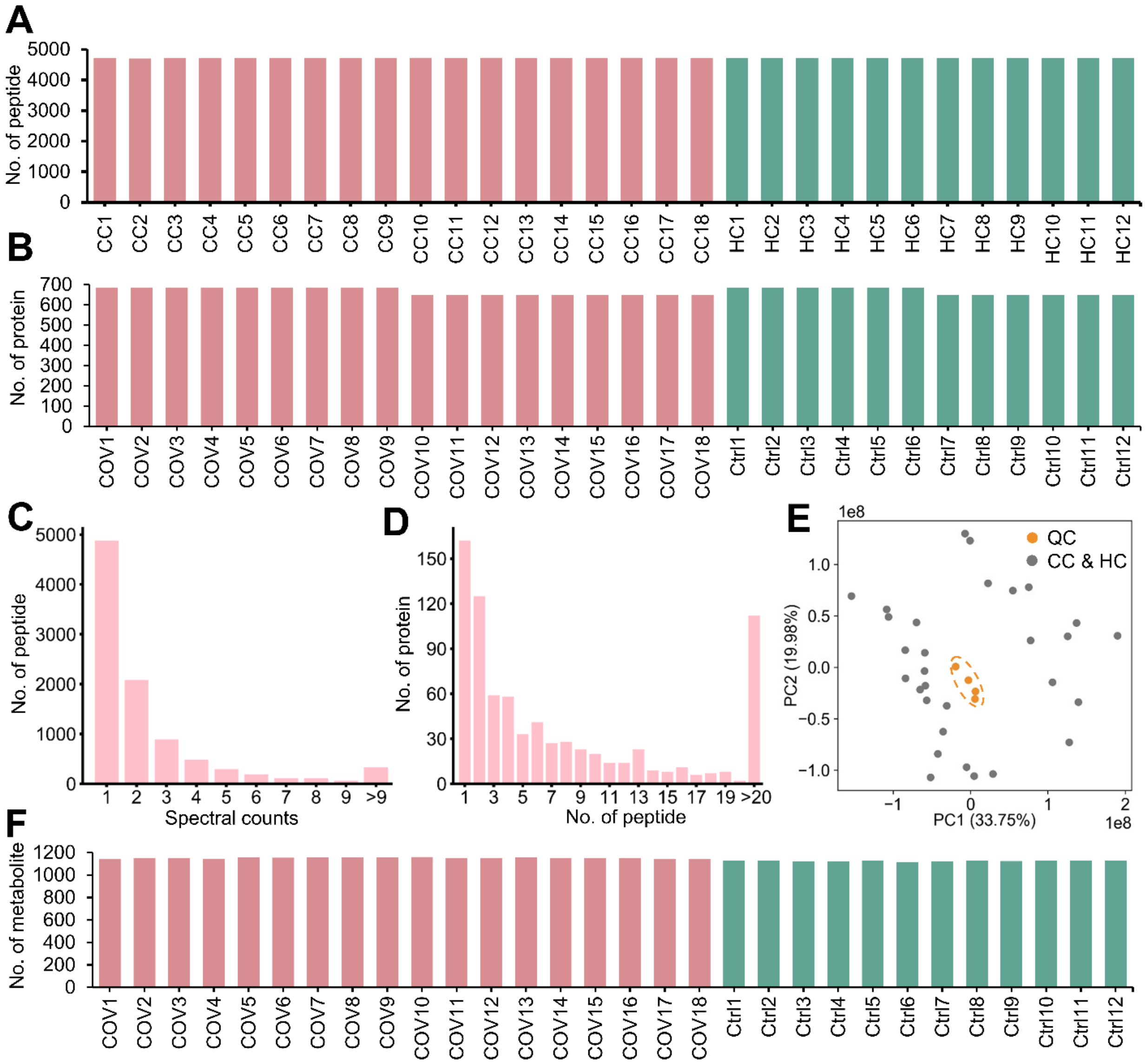


**Fig. S1.**

**Proteomic and metabolomic profiling of plasma from CC and HC cases. A,B** The distribution of numbers of quantified (a) peptides and (b) proteins in the 30 plasma samples. **C** The distribution of MS/MS spectral counts of quantified peptides. **D** The distribution of peptide numbers of quantified proteins. **E** PCA analysis of the 30 metabolomic data and 4 quality controls (QCs). We pipette 10 μL of each sample to pool a QC sample. **F** The distribution of MS/MS spectral counts of quantified metabolites.

Figure S2

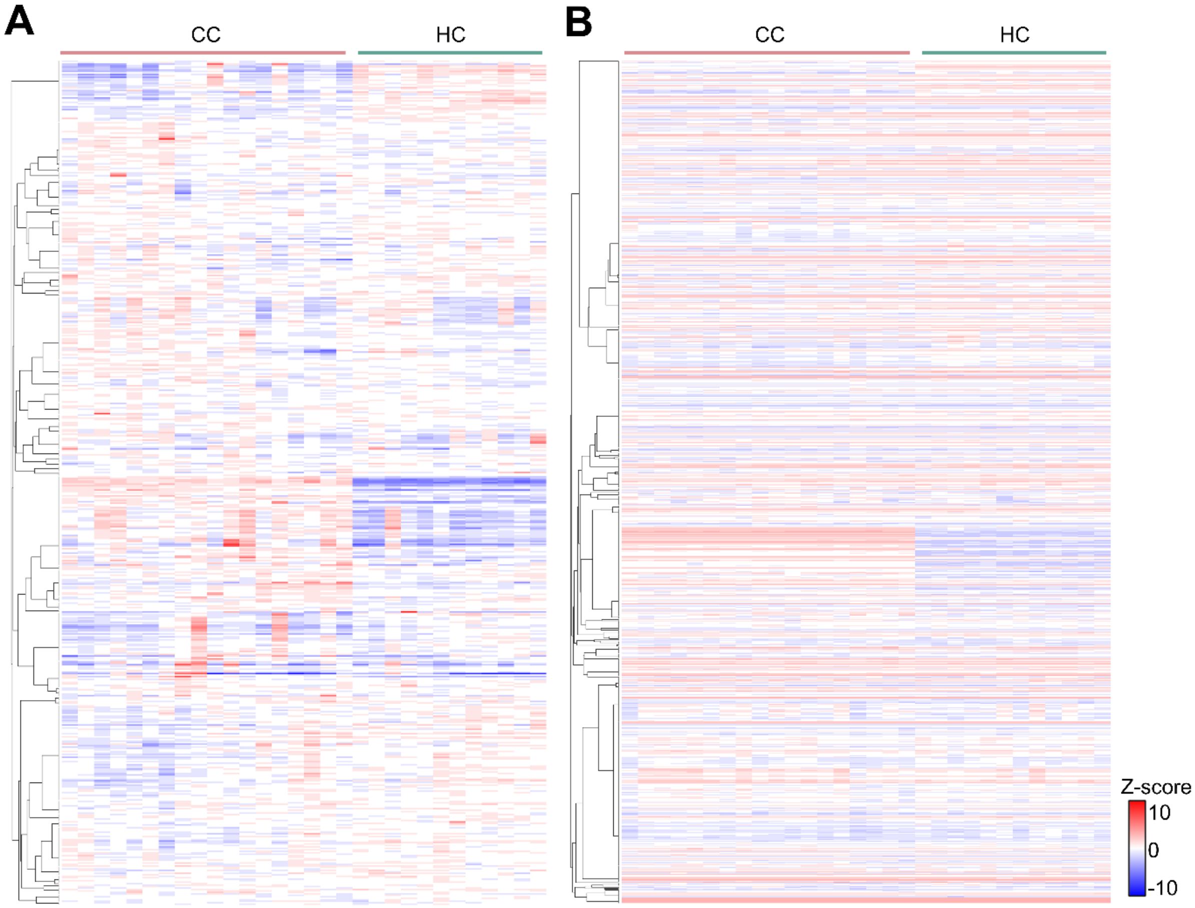


**Fig. S2.**

**The heatmaps for the identified plasma proteins and metabolites**. **A-B** The heatmaps of the finally reserved proteins (A) and metabolites (B).

**Figure S3**

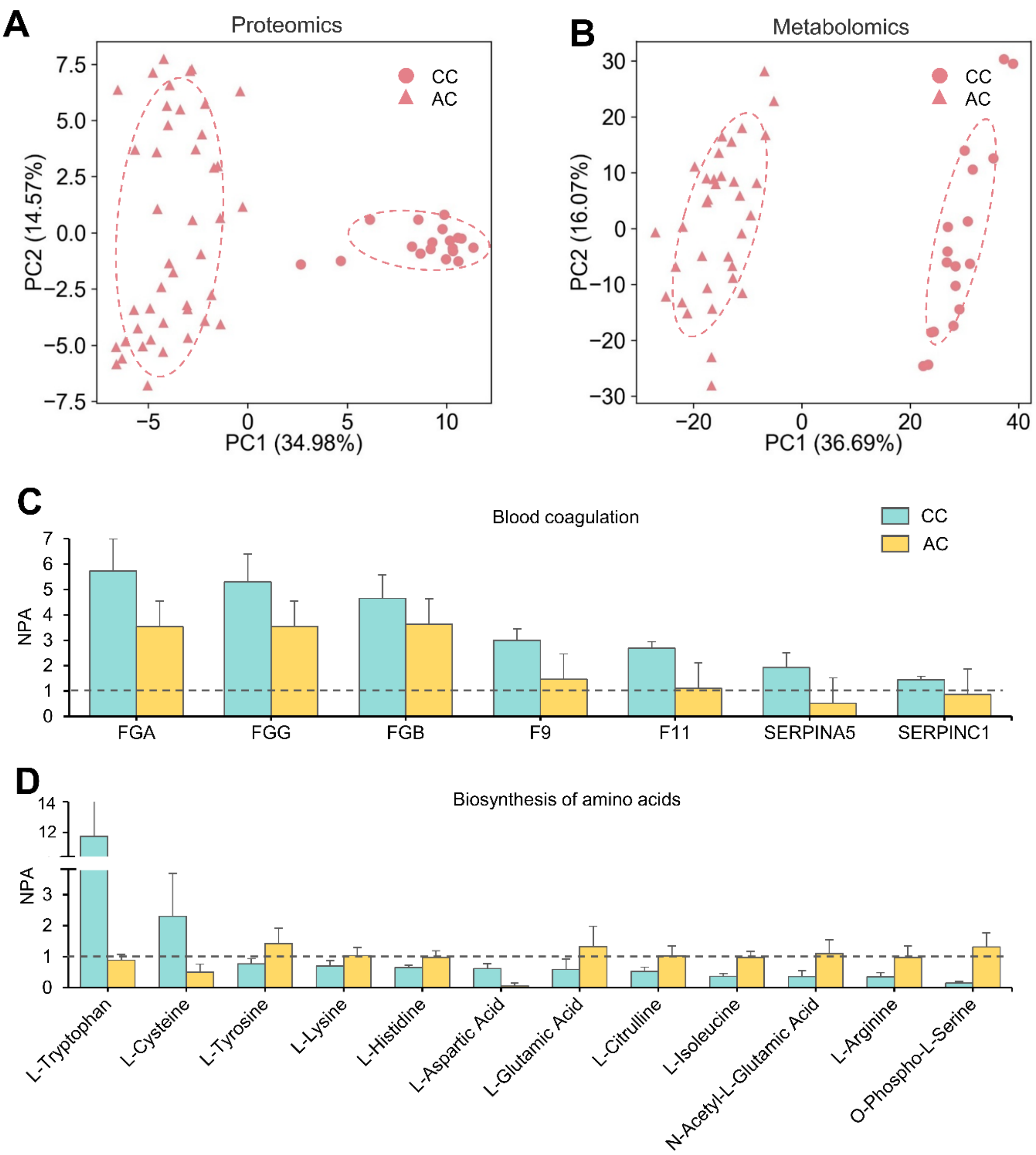


**Fig. S3.**

**The alternations of molecules in CC cases against AC cases. A-B** PCA analyses of the proteomic data of CC and AC (A), and the metabolomics data of CC and AC (B). **C** The abundance of proteins enriched in the process of blood coagulation in CC and AC. **D** The abundance of metabolites enriched in the process of biosynthesis of amino acids in CC and AC.Type or paste caption here. Create a page break and paste in the Figure above the caption.

### Figure S4

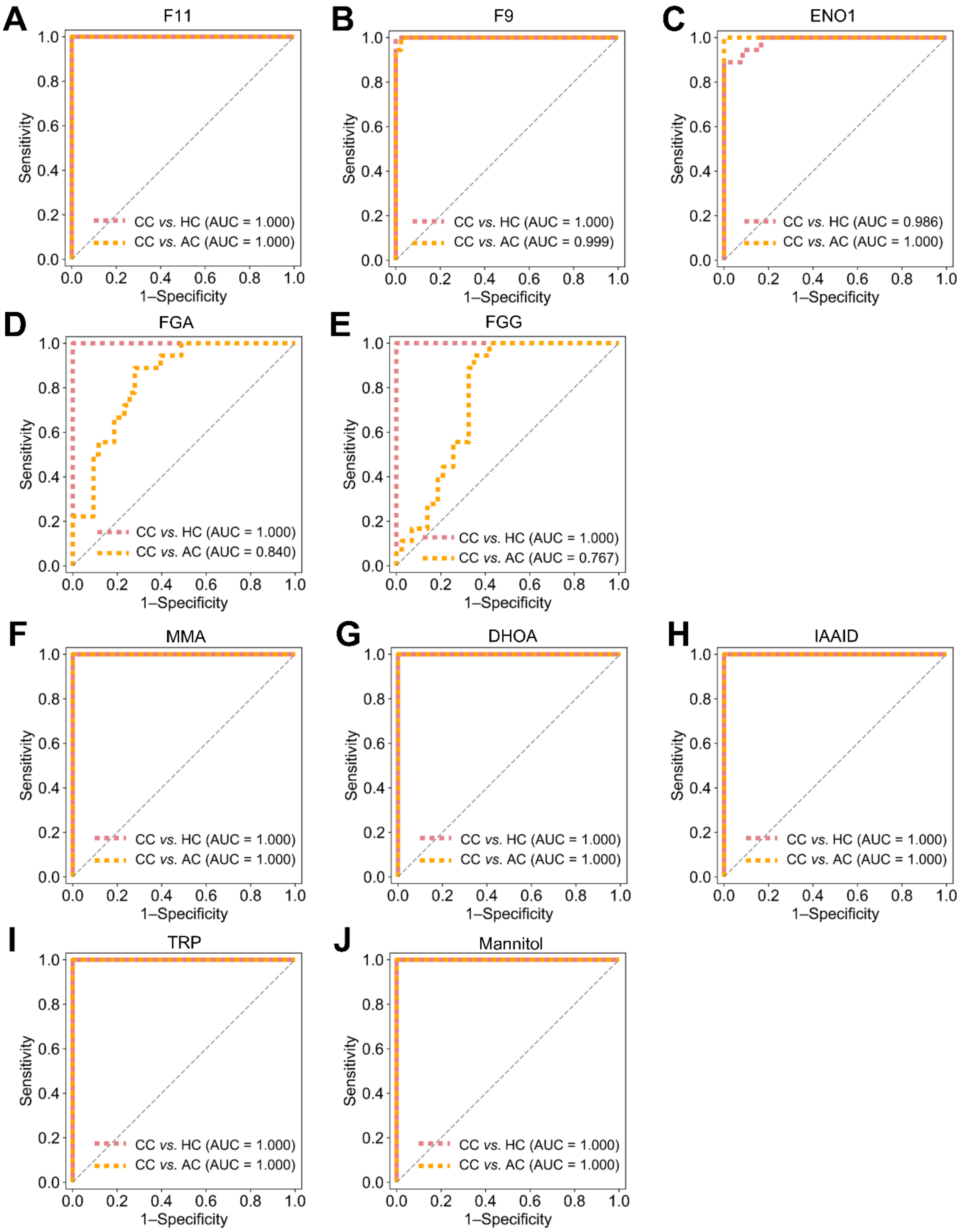
A
F11
Sensitivity
1–Specificity
CC vs. HC (AUC = 1.000)
CC vs. AC (AUC = 1.000)
B
F9
Sensitivity
1–Specificity
CC vs. HC (AUC = 1.000)
CC vs. AC (AUC = 0.999)
C
ENO1
Sensitivity
1–Specificity
CC vs. HC (AUC = 0.986)
CC vs. AC (AUC = 1.000)
D
FGA
Sensitivity
1–Specificity
CC vs. HC (AUC = 1.000)
CC vs. AC (AUC = 0.840)
E
FGG
Sensitivity
1–Specificity
CC vs. HC (AUC = 1.000)
CC vs. AC (AUC = 0.767)
F
MMA
Sensitivity
1–Specificity
CC vs. HC (AUC = 1.000)
CC vs. AC (AUC = 1.000)
G
DHOA
Sensitivity
1–Specificity
CC vs. HC (AUC = 1.000)
CC vs. AC (AUC = 1.000)
H
IAAID
Sensitivity
1–Specificity
CC vs. HC (AUC = 1.000)
CC vs. AC (AUC = 1.000)
I
TRP
Sensitivity
1–Specificity
CC vs. HC (AUC = 1.000)
CC vs. AC (AUC = 1.000)
J
Mannitol
Sensitivity
1–Specificity
CC vs. HC (AUC = 1.000)
CC vs. AC (AUC = 1.000)

**Fig. S4.**

**Individual biomarkers for classifying CC from AC and HC.** The ROC curve and 5-fold cross-validation AUC values of the indicated molecules to classifier the CC cases from HC and AC cases.
